# Epidemiological Profile and Transmission Dynamics of COVID-19 in the Philippines

**DOI:** 10.1101/2020.07.15.20154336

**Authors:** N. J. L. Haw, J. Uy, K. T. L. Sy, M. R. M. Abrigo

**Affiliations:** Health Sciences Program, School of Science and Engineering, Ateneo de Manila University; Philippine Institute for Development Studies; Department of Epidemiology, Boston University School of Public Health; Department of Global Health, Boston University School of Public Health

**Author notes:** Corresponding author details N. J. L. HAW, Health Sciences Program, School of Science and Engineering, Ateneo de Manila University, Katipunan Avenue, Loyola Heights, Quezon City, Philippines 1108, local 5618. Co-first authors: Nel Jason L. Haw and Jhanna Uy contributed equally to this article.

## Abstract

The Philippines confirmed local transmission of COVID-19 on 7 March 2020. We described the characteristics and epidemiological time-to-event distributions for laboratory-confirmed cases in the Philippines. The median age of 8,212 cases was 46 years (IQR: 32-61), with 46.2% being female and 68.8% living in the National Capital Region. Health care workers represented 24.7% of all detected infections. Mean length of hospitalization for those who were discharged or died were 16.00 days (95% CI: 15.48, 16.54) and 7.27 days (95% CI: 6.59, 8.24). Mean duration of illness was 26.66 days (95% CI: 26.06, 27.28) and 12.61 days (95% CI: 11.88, 13.37) for those who recovered or died. Mean serial interval was 6.90 days (95% CI: 5.81, 8.41). Epidemic doubling time pre-quarantine (11 February and 19 March) was 4.86 days (95% CI: 4.67, 5.07) and the reproductive number was 2.41 (95% CI: 2.33, 2.48). During quarantine (March 20 to April 9), doubling time was 12.97 days (95% CI: 12.57, 13.39) and the reproductive number was 0.89 (95% CI: 0.78, 1.02).

## INTRODUCTION

Coronavirus Disease 2019 (COVID-19) was declared a global pandemic by the World Health Organization (WHO) on 12 March 2020.[1] Current and published epidemiological research on COVID-19 have largely focused on China and other high-income countries such as South Korea, Japan, the United States, Italy, and Spain.[2] Further research on the distribution and burden of COVID-19 in low- and middle-income countries (LMICs) may give insight on its disease epidemiology in low resource settings as transmission dynamics are dependent not only on population characteristics,[3] but also health system capacity (e.g. access to testing),[4] and the ability to implement mitigation measures (e.g. community-level quarantine, social distancing).[5] We describe the epidemiological profile and transmission dynamics of the first 8,212 confirmed COVID-19 cases in the Philippines.

## METHODS

### Overview of the Philippine COVID-19 Surveillance System

The Philippines is an archipelago of three island groups and 17 regions subdivided into 81 provinces covering 146 cities and 1,488 municipalities.[6] COVID-19 surveillance, like majority of health service delivery, is decentralized to local government units (LGUs), i.e. provinces, cities, and municipalities. Epidemiology and surveillance units (ESUs) exist in every administrative level, namely regional ESUs (RESUs), provincial ESUs (PESUs), and city/municipality ESUs (CESUs/MESUs). Units collect data for their jurisdictions and report to higher level units: CESUs/MESUs cascade daily updates to the PESUs, which cascade those to regional ESUs which then finally submit to the DOH-Epidemiology Bureau (EB).[7,8] The 15 cities and one municipality in the National Capital Region (NCR), as well as 37 highly urbanized cities (HUCs) and independent component cities (ICCs) are not overseen by a provincial government, and report directly to the RESUs.

The DOH-EB is the lead national agency for COVID-19 surveillance. It collates data on confirmed and suspected cases nationwide and provides guidance and support to all LGUs. DOH-EB maintains an information system for COVID-19 cases patterned after influenza-like illness (ILI)/severe acute respiratory infection (SARI) surveillance. Confirmed cases are profiled using case investigation forms (CIF), which record patient characteristics, epidemiologic links, and select clinical information.

### Case Definition, Case Detection, and Laboratory Testing

We define COVID-19 cases as patients with positive real-time reverse transcription polymerase chain reaction (RT-PCR) conducted by laboratories accredited by the DOH and Research Institute for Tropical Medicine (RITM).[9,10] The RITM is the National Reference Laboratory for Emerging and Re-emerging Diseases and it is the public health authority that accredits laboratories for COVID-19 testing. We included cases recorded up to 29 April 2020 at the time of writing and followed the status of these patients until 22 May 2020 using the latest data set prior to publication.

Starting 9 April 2020, the DOH limits testing to suspect and probable cases. A suspect case is a person who has any of the following: (1) SARI requiring hospitalization with no other etiology that fully explains clinical presentation; (2) ILI with no other etiology that fully explains clinical presentation AND residence or travel to an area with known local transmission 14 days prior to symptoms OR exposure to confirmed or probable cases during the period two days prior to symptoms until they test negative with RT-PCR; and (3) high-risk groups presenting with fever, cough, shortness of breath, and other respiratory symptoms, including the elderly 60 years and above, those with comorbidities, women with high-risk pregnancies, and health workers.[11] Probable cases are suspect cases (1) referred for RT-PCR testing, (2) with inconclusive RT-PCR results from a DOH-accredited laboratory, or (3) who have a positive RT-PCR result from a non-RITM accredited laboratory. As of 29 April, there have been six versions of COVID-19 case definitions and testing has expanded to include 17 subnational laboratories (see Supplementary Appendix).

Potential cases are detected through multiple avenues. At ports of entry, the DOH - Bureau of Quarantine identifies persons showing symptoms such as fever, shortness of breath, and respiratory problems and refers them to health facilities, LGU health offices, or sentinel disease reporting units (DRUs). Health providers also use official case definitions to assess potential COVID-19 cases among patients who consult or are hospitalized in their facilities. LGUs, ESUs, and DRUs then conduct case investigations and contact tracing for reported confirmed, suspect, and probable COVID-19 cases.[11]

### Statistical Analysis

We analyzed descriptive statistics of cases, deaths, and recoveries by socio-demographics, symptoms at specimen collection, and health care worker status to the extent that these data were available. Epidemic curves were constructed using the date of symptom onset. As many dates were missing, they were imputed based on the method by Günther et al. as applied to the COVID-19 outbreak in Bavaria, Germany.[12] A flexible, generalized additive model with a Weibull distribution was fitted for the days between symptom onset and DOH public announcement. Predictors were: (1) day of week of reporting, (2) region of residence, (3) laboratory of testing, (4) sex; and the following smoothed predictors: (5) calendar week of reporting date; and (6) age (Supplementary Appendix). The imputed data set was used for all further analyses.

Among a subset of cases with complete data, we estimated the distributions of six time-to-event variables: (1) serial interval, or time between symptom onset of index and secondary cases; (2) health-seeking behavioural delays, or time between symptom onset and first medical consultation (or specimen collection if missing); (3) diagnostic delays, or time between specimen collection (or first medical consultation if missing) and laboratory confirmation; (4) hospital length of stay for admitted cases, or time between admission and discharge (or death for those who died); (5) length of condition, or time between symptom onset and death or recovery; (6) reporting delays for confirmed cases, or time between laboratory confirmation and public announcement. To calculate the serial interval, clustering analysis was done using known epidemiologic links, exposure history, and symptom onset for the first 100 confirmed COVID-19 cases where contact tracing was mandated by DOH guidelines.[13] Distributions that best fit the data were identified using the Akaike information criterion (AIC).

Exponential growth rates, epidemic doubling times, and reproductive numbers (basic reproductive number (R0) and effective reproductive number (Re) were calculated, accounting for the empirical serial interval of our data.[14,15] R0 was estimated during the exponential growth period using the method developed by Wallinga and Lipsitch,[16] with the best fit of dates determined by the deviance based *R*^2^ statistic.[17] Re was estimated using the Wallinga and Teunis method during the period after community quarantine was mandated.[18]

Analyses and visualizations were done using Stata 16.1 (College Station, TX: StataCorp LP), Tableau Desktop 2020.1.3 (Mountain View, CA: Tableau Software, Inc), and R 4.0.0[19] using the EpiEstim,[20] epitools,[21] fitdistrplus,[22] gamlss,[23] and MASS[24] packages.

#### Ethics Approval

Data came from continuing surveillance efforts by the DOH-EB as part of an ongoing outbreak investigation. Thus, institutional review board approval was deemed unnecessary.

## RESULTS

Among the 8,212 COVID-19 cases detected in the Philippines up to 29 April 2020, 46.2% were female and 68.8% lived in the NCR (Table 1). Among these, 768 (9.4%) died and 2,988 (36.4%) recovered. Median age for cases, deaths, and recoveries were 46 years (IQR: 32-61), 66 years (IQR: 57-74), and 46 years (IQR: 32-59). There were 319 (3.9%) cases and 12 deaths among those aged 0 to 20 years. Health care workers represent 24.7% of detected infections with 35 deaths. Hypertension (17.9%) and Diabetes (12.7%) were the most common comorbidities while fever (31.3%) and cough (44.9%) were the most common symptoms at specimen collection. Compared to those who recovered, those who died were more likely to be older, male, presented with difficulty breathing, and had comorbidities except for asthma.

**Table 1.**
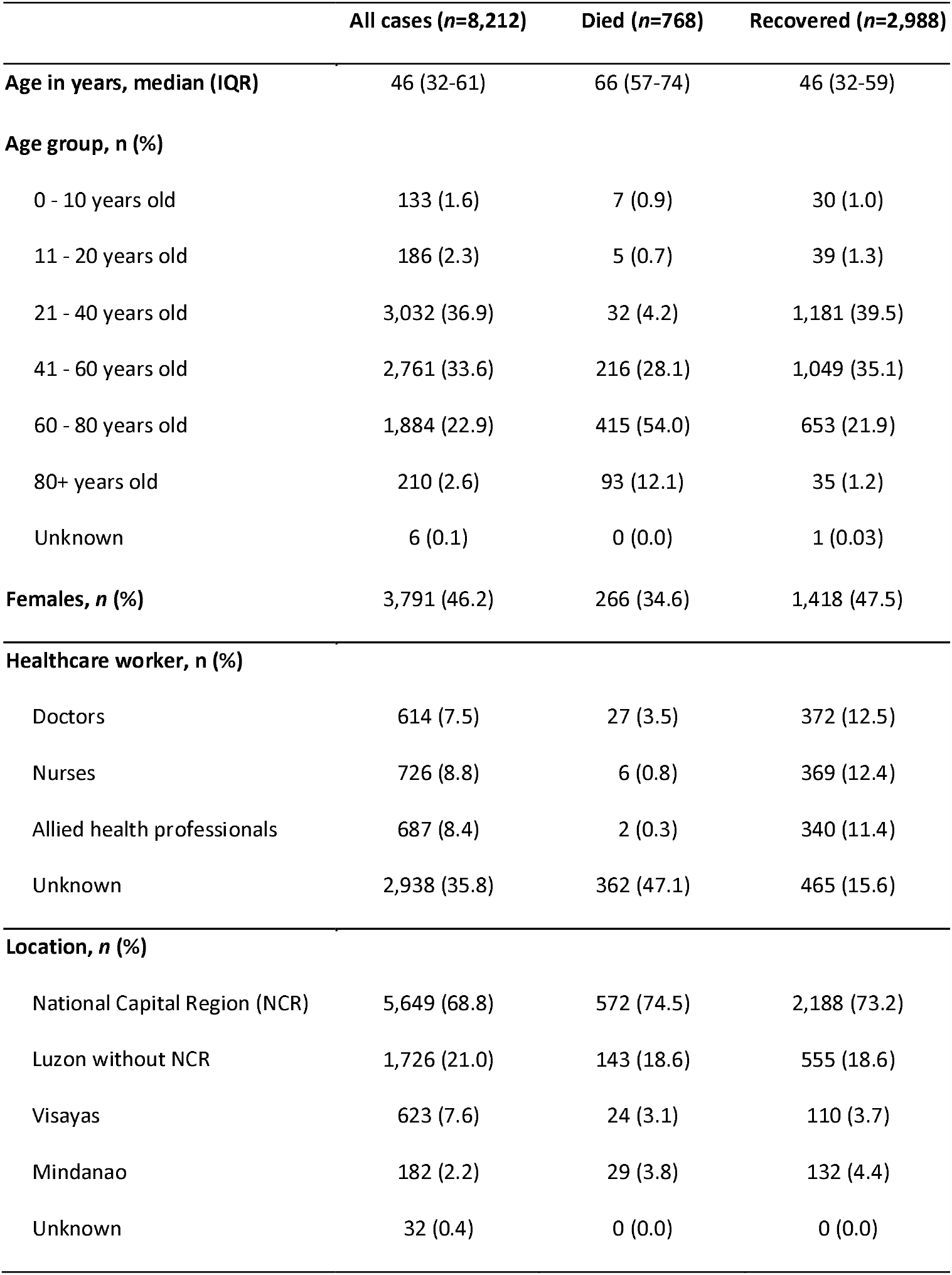
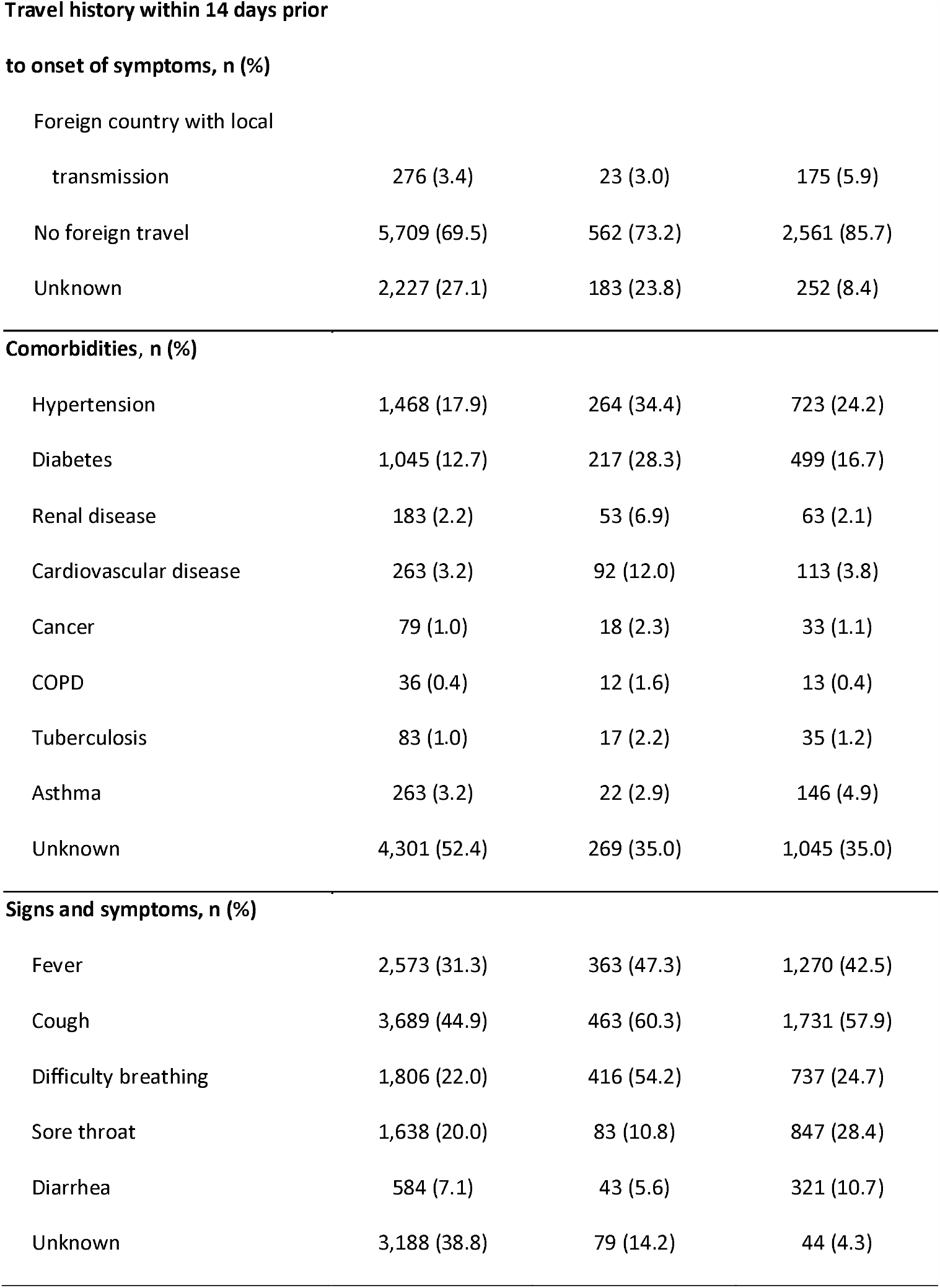
Characteristics of COVID-19 cases in the Philippines as of 29 April 2020.

The epidemic curves based on symptom onset shows that the exponential growth period of the outbreak likely began on 11 February 2020 (Figure 1). Only 5,168 (63.0%) cases had complete dates of symptom onset and the rest were imputed (see Supplementary Appendix for model diagnostics). Several cases (3.5%) visited a country with known local transmission 14 days prior to their symptoms. New cases steadily increased until the week of 15 March 2020 when NCR and Luzon were put under strict community-level quarantine. Detected cases rose during this period likely due to the expansion of laboratory testing capacity in the last week of March. The decreases in the last weeks leading up to 29 April were likely due to delays between symptom onset and public announcement - and not due to a true decline in new infections.

**Figure 1.**
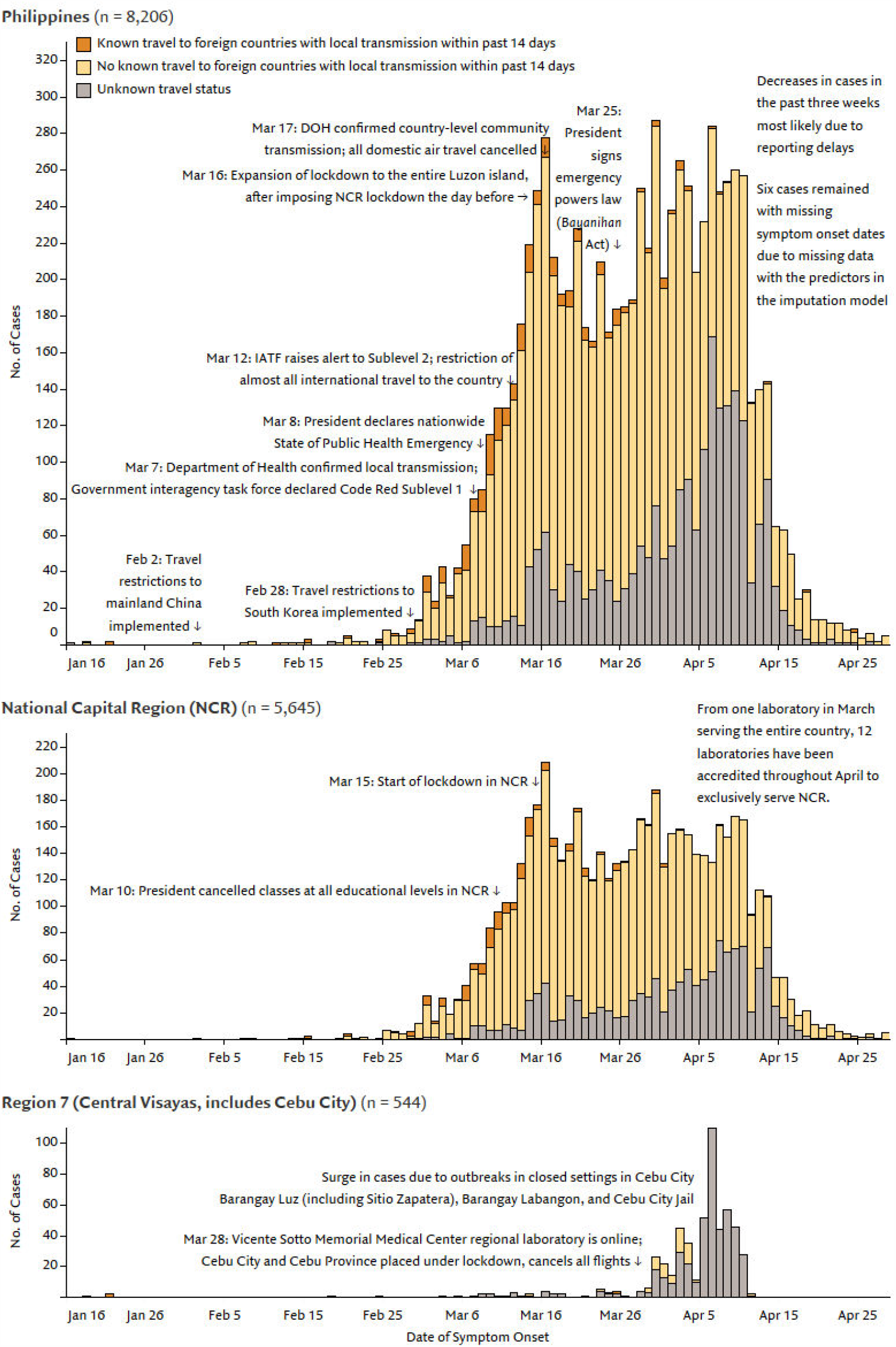
Onset of illness among the first 8,212 COVID-19 cases in the Philippines, the National Capital Region, and Region 7

Figure 2 and Supplementary Appendix presents the distributions for six key epidemiologic events and surveillance delays from symptom onset to public announcement in the Philippine health system. The serial interval had a mean of 6.90 days (95% CI: 5.81, 8.41; Weibull) using 55 pairs of index and secondary cases. The mean health-seeking behavioural delay or time between illness onset and first medical consultation was 6.75 days (95% CI: 6.70, 7.15; gamma). The mean diagnostic delay or time between specimen collection to laboratory confirmation was 4.92 days (95% CI 4.82, 5.02; lognormal). Among those who were hospitalized, the mean length of stay for recoveries was 16.00 days (95% CI: 15.48, 16.54; Weibull) while for deaths was 7.27 days (95% CI: 6.59, 8.24; gamma). Among those who had recovered or died, the mean duration of illness with COVID-19 was 26.66 days (95% CI: 26.06, 27.28; Weibull) and 12.61 days (95% CI: 11.88, 13.37; gamma), respectively. Lastly, mean reporting delays or the time between laboratory confirmation and public announcement was 2.47 (95% CI: 2.40, 2.51; gamma). Notably, these delays were such that of the 541 deaths with complete data on date of death and laboratory confirmation, half of them (276, 51%) received laboratory confirmation after they had died.

**Figure 2.**
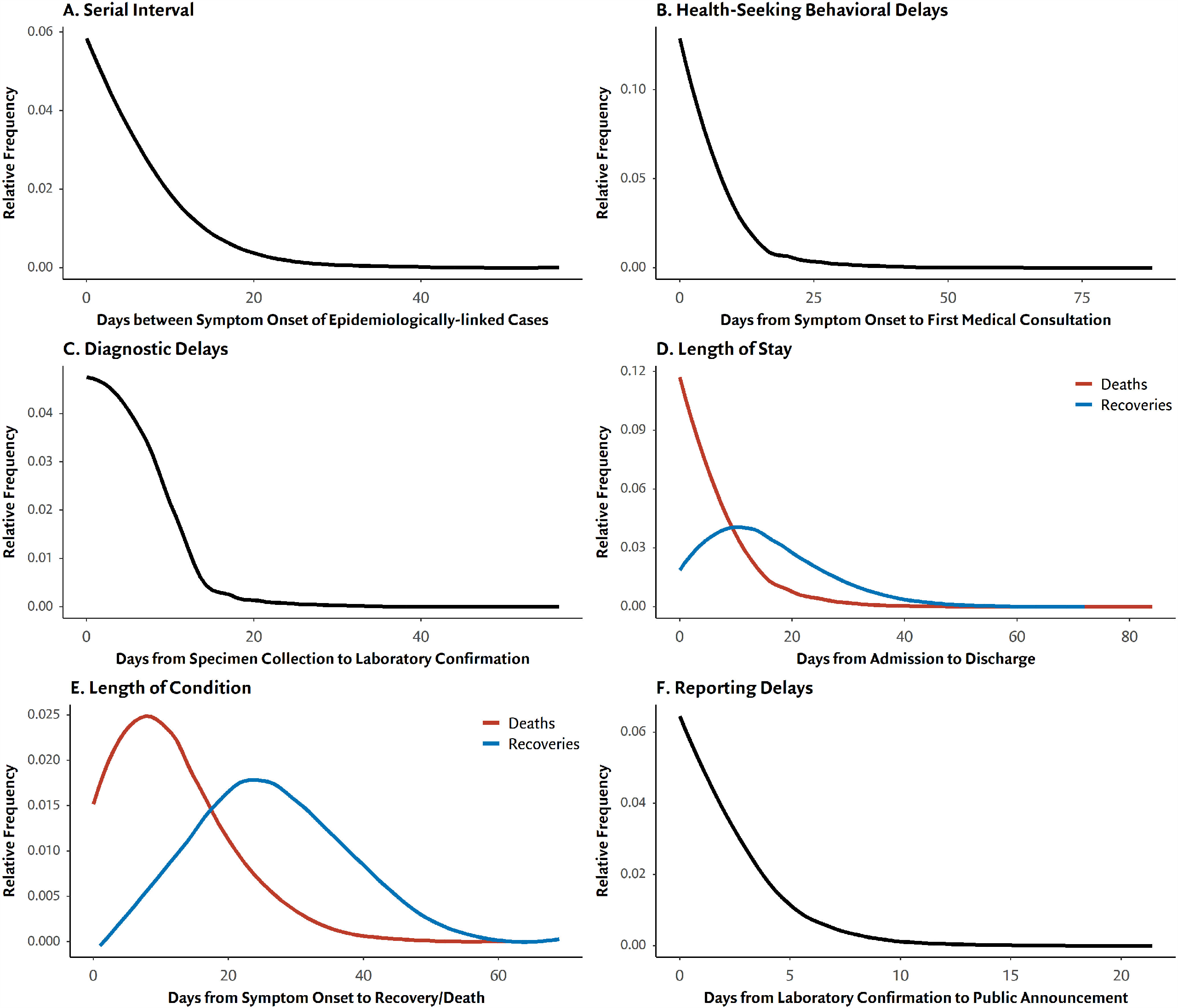
Time-to-Event Distributions for key epidemiologic events and surveillance delays A) Serial interval or the time between onset of symptoms for index and secondary cases B) Behavioral delay or time between symptom onset to first medical consultation C) Diagnostic delay or time between illness onset to laboratory confirmation for COVID-19 D) Length of stay or time from hospitalization to discharge (blue) or death (red) E) Length of condition or time from symptom onset to recovery (blue) or death (red) F) Reporting delay or time between laboratory confirmation to public announcement by DOH

Figure 3 presents three of the largest known clusters. Cluster A’s likely source of infection was the wake of a cases’ mother-in-law; it was also plausible the first generation of cases had multiple points of contact before and after the wake given that they were all friends and family. The second-generation cases were household members of first generation cases, save for one who infected a high-level government official. Cluster B and C’s likely sources of infection were birthday parties, and similar to Cluster A, infected the rest of their household. Aside from friends and family, other known epidemiological links are between cases and their attending health care providers, such as the one found in Cluster A.

**Figure 3.**
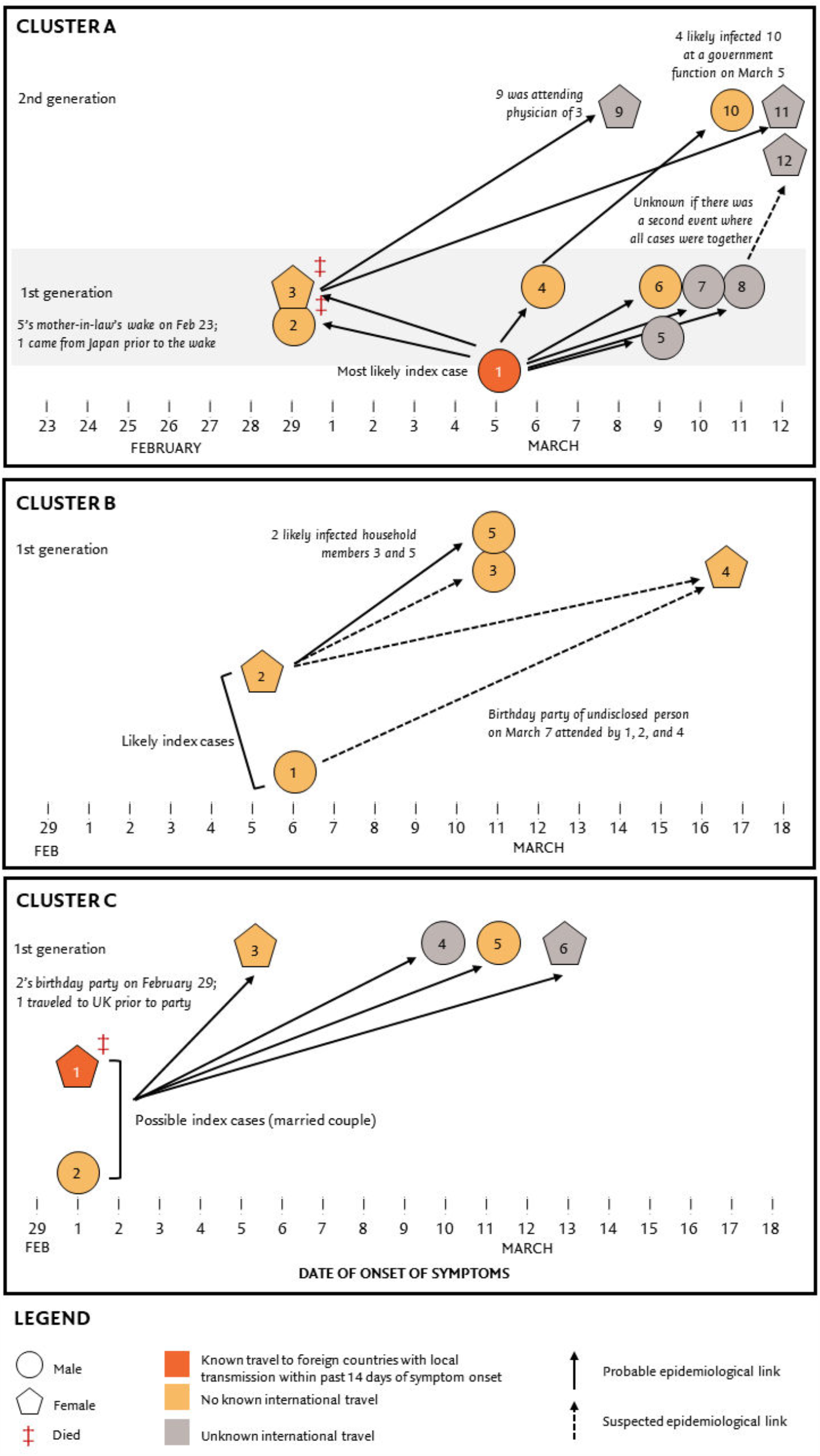
Some known large Covid-19 outbreak clusters in the Philippines

In the exponential growth phase pre-quarantine (February 11 and March 19), the epidemic growth rate was 0.143 per day (95% CI: 0.137, 0.149), and the epidemic doubling time was 4.86 days (95% CI: 4.67, 5.07). Using the estimated serial interval distribution, the basic reproductive number for this period was 2.41 (95% CI: 2.33, 2.48). Analysis for the quarantine period was limited to April 15 to account for reporting delays. In the period of March 20 to April 15, the epidemic growth rate was 0.053 per day (95% CI: 0.052, 0.055), the epidemic doubling time was 12.97 days (95% CI: 12.57, 13.39), and the average daily effective reproductive number was 0.89 (95% CI: 0.78, 1.02).

## DISCUSSION

This overview of the first three months of the COVID-19 outbreak in the Philippines provides a unique perspective on the transmission of COVID-19 in a health system with limitations in surveillance, testing, and service delivery. Such information may aid modelling or data analyses efforts for outbreak response in the Philippines and countries with similar health system constraints as studies of the pandemic outside of high-income countries and in LMICs have been limited.

Our results support findings that COVID-19 disproportionately burdens older age groups, healthcare workers, and densely populated urban areas.[25–27] Outbreak epicentres in the Philippines were urban centres, such as NCR and Cebu City, where NCR alone accounted for more than two-thirds of all cases. NCR is the fifth most densely populated metropolis in the world.[28] The first few cases were likely imported into these urban centres, as approximately one-tenth of the population are migrant workers and 8 million tourists visit annually.[29] The high proportion of healthcare workers as a share of cases may indicate poor compliance to infection control protocols and inadequate availability of personal protective equipment. In the Philippines, one in four confirmed cases were healthcare workers, much higher than the Western Pacific regional average of 2-3%.[30] Further investigation is necessary to identify exposure sources, gaps in infection control, and facilitators of hospital outbreaks.

In the Philippines, surveillance delays were prominent due to the limited healthcare system capacity which affects the timeliness of decisions to suppress the outbreak. For example, on 7 March, the DOH officially reported local transmission upon public announcement of the sixth case,[31,32] but our findings suggest that the exponential growth period likely began more than three weeks earlier. Delays also affect rapid isolation of suspected cases, which is critical to reduce the reproductive number to below 1 and suppress the outbreak. To reduce health seeking behavioural delays, the National Health Insurance Program, of which all Filipinos are members under the newly passed Universal Health Care Law, has made laboratory testing free[33] and covers the majority of community isolation and hospitalization costs since mid-April.[34–36] To reduce diagnostic delays, the Philippines has slowly expanded laboratory capacity from just one laboratory in February. More isolation facilities, such as stadiums, hotels, and schools, have been set-up for mild cases to save limited hospital resources for severe and critical cases. Information systems are being strengthened to allow synchronized reporting of cases at all ESU levels in real time.

Our analysis also showed that the pre-quarantine reproductive number was 2.4, meaning that for every confirmed case, between two and three others cases were infected. This estimate is similar to previous estimates in literature.[37] Travel restrictions imposed for China and South Korea in February may not have been sufficient to slow the progression of the outbreak, as the reproductive number remained around 2.4 during the period before community-level quarantines were implemented. While full travel restrictions and community-level quarantines decreased the reproductive number to 0.9, these interventions merely delayed the progression of the outbreak rather than stopping it completely. Furthermore, the serial interval estimate of 7 days is longer but comparable to previous estimates in other settings.[38]

This study has limitations common in disease outbreak response and health information systems in LMICs. First, there is a large number of missing data that needs to be further validated due to difficulties of retrieving data from a decentralized surveillance system. Data were from paper case investigation forms and not medical records. As such, we did not have detailed clinical information on disease severity or disease progression, and comorbidities may be underestimated. Only 63% of cases reported their date of symptom onset; however, we implemented a previously validated imputation method to correctly specify the epidemic curve and better estimate the reproductive number. Second, the focus of laboratory testing for early phases of the Philippine outbreak were on severe or critical cases, but the DOH has since expanded testing for those with milder symptoms. Since more milder cases were captured by the surveillance system during the latter part of the outbreak, mean surveillance delays, length of stay, and duration of illness may have declined as health system capacity improves. Thus, our estimates of the epidemiologic events and surveillance delays may be overestimated. In addition, the case counts may be artificially undercounted due to underreporting of mild cases, missed asymptomatic cases, and backlogs in confirmatory testing. Thus, we did not calculate the case fatality rate due to these potential biases. Finally, there may be additional differential ascertainment in the characteristics of cases due to disparities in access to care; individuals with higher socioeconomic status, health care workers, and workers with occupations who are able to stay sick at home may be more likely to test and seek care, even though socially disadvantaged communities may have increased susceptibility to COVID-19.[39]

## Data Availability

Most data are publicly available at the Department of Health Philippines Official COVID-19 tracker

https://ncovtracker.doh.gov.ph

## REFERENCES

1. World Health Organization. Coronavirus disease 2019 (COVID-19) Situation Report – 52. WHO, 2020.

2. World Health Organization. Global research on coronavirus disease (COVID-19). 2020(https://www.who.int/emergencies/diseases/novel-coronavirus-2019/global-research-on-novel-coronavirus-2019-ncov).

3. House T, Keeling MJ. Household structure and infectious disease transmission. Epidemiology & Infection Cambridge University Press, 2009; 137: 654–661.

4. Kelly-Cirino CD, et al. Importance of diagnostics in epidemic and pandemic preparedness. BMJ Global Health BMJ Specialist Journals, 2019; 4: e001179.

5. Abrigo MRM, et al.Projected Disease Transmission, Health System Requirements, and Macroeconomic Impacts of the Coronavirus Disease 2019 (COVID-19) in the Philippines. Philippines: Philippine Institute for Development Studies, 2020 Apr. Report No.: No. 2020-015.

6. Philippine Statistics Authority. Philippine Standard Geographic Codes as of 31 December 2019. 2019.

7. Department of Health. Department Memorandum 2020-0115: Prioritization of Contact Tracing for Confirmed Coronavirus Disease 2019 (COVID-19) Cases. 2020.

8. Seventeenth Congress of the Philippines. Mandatory Reporting of Notifiable Diseases and Health Events of Public Health Concern Act (Republic Act 11332).

9. Food and Drug Administration Philippines. List of Approved COVID-19 Test Kits for Commercial Use (as of 26 Mar 2020, 2:00 PM). 2020.

10. Department of Health. Department Circular 2020-0143: Public Advisory No. 20: COVID-19 Laboratory Testing. 2020.

11. Department of Health. Administrative Order No. 2020-0013: Revised Administrative Order No. 2020-0012 ‘Guidelines for the Inclusion of the Coronavirus Disease 2019 (COVID-19) in the List of Notifiable Diseases for Mandatory Reporting to the Department of Health’ dated March 17, 2020. 2020.

12. Günther F, et al.Nowcasting the COVID-19 Pandemic in Bavaria. Munich, Germany: Statistical Consulting Unit StaBLab, 2020 Apr.

13. Department of Health. Department Memorandum No. 2020-0115: Prioritization of Contact Tracing for Confirmed Coronavirus Diseases 2019 (COVID-19) Cases. 2020.

14. Li Q, et al. Early Transmission Dynamics in Wuhan, China, of Novel Coronavirus–Infected Pneumonia. New England Journal of Medicine Massachusetts Medical Society, 2020; 382: 1199–1207.

15. Lauer SA, et al. The Incubation Period of Coronavirus Disease 2019 (COVID-19) From Publicly Reported Confirmed Cases: Estimation and Application. Annals of Internal Medicine 2020; Published online: 10 March 2020.doi:10.7326/M20-0504.

16. Wallinga J, Lipsitch M. How generation intervals shape the relationship between growth rates and reproductive numbers. Proceedings of the Royal Society B: Biological Sciences Royal Society, 2007; 274: 599–604.

17. Obadia T, Haneef R, Boëlle P-Y. The R0 package: a toolbox to estimate reproduction numbers for epidemic outbreaks. BMC Medical Informatics and Decision Making 2012; 12: 147.

18. Wallinga J, Teunis P. Different epidemic curves for severe acute respiratory syndrome reveal similar impacts of control measures. American Journal of Epidemiology 2004; 160: 509–516.

19. R Core Team. R: A language and environment for statistical computing. Vienna, Austria: R Foundation for Statistial Computing, 2020.

20. Cori A. EpiEstim: Estimate Time Varying Reproduction Numbers from Epidemic Curves. 2019.

21. Aragon TJ. epitools: Epidemiology Tools. 2020.

22. Delignette-Muller ML, Dutang C. fitdistrplus: An R Package for Fitting Distributions. Jouranl of Statistical Software 2015; 64: 1–34.

23. Rigby RA, Stasinopoulos DM. Generalized additive models for location, scale and shape. Journal of the Royal Statistical Society: Series C (Applied Statistics) 2005; 54: 507–554.

24. Venables WN, Ripley BD. Modern applied statistics with S. 4th ed. New York: Springer, 2002.

25. Wu Z, McGoogan JM. Characteristics of and Important Lessons From the Coronavirus Disease2019 (COVID-19) Outbreak in China: Summary of a Report of 72 314 Cases From the Chinese Center for Disease Control and Prevention. JAMA American Medical Association, 2020; 323: 1239–1242.

26. Bi Q, et al. Epidemiology and transmission of COVID-19 in 391 cases and 1286 of their close contacts in Shenzhen, China: a retrospective cohort study. The Lancet Infectious Diseases Elsevier, 2020; 0Published online: 27 April 2020.doi:10.1016/S1473-3099(20)30287-5.

27. CDCMMWR. Characteristics of Health Care Personnel with COVID-19 — United States, February 12–April 9, 2020. MMWR. Morbidity and Mortality Weekly Report 2020; 69Published online: 2020.doi:10.15585/mmwr.mm6915e6.

28. Demographia. Demographia World Urban Areas. 16th ed. Illinois, USA: Demographia, 2020. 29. Department of Tourism. Tourism Statistics 2019. Makati City: DOT, 2020.

29. Esguerra CV. Health Workers in Philippines catching COVID-19 ‘worrisome’: WHO official. ABS-CBN News. Manila, 2020; Published online: 21 April 2020.

30. Department of Health. DOH CONFIRMS LOCAL TRANSMISSION OF COVID-19 IN PH; REPORTS 6TH CASE. Department of Health. (https://www.doh.gov.ph/doh-press-release/doh-confirms-local-transmission-of-covid-19-in-ph). Accessed 22 April 2020.

31. Duterte RR. Proclamation No. 922. Declaring a State of Public Health Emergency throughout the Philippines. Republic of the Philippines, 2020.

32. Philippine Health Insurance Corporation. Benefit package for testing for SARS-CoV-2. Pasig City, Philippines: PHIC, 2020 Apr. Report No.: 2020–0010.

33. Philippine Health Insurance Corporation. Guidelines on the COVID-19 Community Isolation Benefit Package (CCIBP). Pasig City, Philippines: PHIC, 2020 Apr. Report No.: 2020–0012.

34. Philippine Health Insurance Corporation. Benefit packages for inpatient care of probable and confirmed COVID-19 developing severe illness/outcomes. Pasig City, Philippines: PHIC, 2020 Apr. Report No.: 2020–0009.

35. Philippine Health Insurance Corporation. Full financial risk protection for Filipino health workers and patients against coronavirus disease (COVID-19). Pasig City, Philippines: PHIC, 2020 Apr. Report No.: 2020–0011.

36. Park M, et al. A Systematic Review of COVID-19 Epidemiology Based on Current Evidence. Journal of Clinical Medicine Multidisciplinary Digital Publishing Institute, 2020; 9: 967.

37. Nishiura H, Linton NM, Akhmetzhanov AR. Serial interval of novel coronavirus (COVID-19) infections. International journal of infectious diseases: IJID: official publication of the International Society for Infectious Diseases 2020; 93: 284–286.

38. Chung RY-N, Dong D, Li MM. Socioeconomic gradient in health and the covid-19 outbreak. BMJ British Medical Journal Publishing Group, 2020; 369Published online: 1 April 2020.doi:10.1136/bmj.m1329.

